# Transparency Assessment of COVID-19 Models

**DOI:** 10.1101/2020.07.18.20156851

**Authors:** Mohammad S. Jalali, Catherine DiGennaro, Devi Sridhar

## Abstract

As the COVID-19 pandemic has caused major societal unrest, modelers have worked to project future trends of COVID-19 and predict upcoming challenges and impacts of policy action. These models, alone or in aggregate, are influential for decision-makers at every level. Therefore, the method and documentation of COVID-19 models must be highly transparent to ensure that projections and consequential policies put forth have sound epistemological grounds. We evaluated 29 COVID-19 models receiving high attention levels within the scientific community and/or informing government responses. We evaluated these models against 27 transparency criteria. We found high levels of transparency in model documentation aspects such as reporting uncertainty analysis; however, about half of the models do not share code and a quarter do not report equations. These discrepancies underscore the need for transparency and reproducibility to be at the forefront of researchers’ priorities, especially during a global health crisis when stakes are critically high.

**Summary:** Evaluation of 29 impactful COVID-19 models reveals inconsistent adherence to best transparency practices; higher transparency is needed to inform policy.

## Main text

The COVID-19 pandemic has strained societal structures and created a global crisis. Scientific models play a critical role in mitigating the pandemic’s harm, from estimating the spread of outbreaks to analyzing the effects of public health policies. Given the gravity of this crisis, the context- and time-sensitive measures with real population health impacts that COVID-19 models provide are of utmost importance. These models must be completely transparent before policies and insights are enacted.

Transparency is the cornerstone of the scientific method and efforts to improve transparency and reproducibility of research have been increasing over the past few years (*1*). Recently, *Science* called for complete transparency of COVID-19 models (*2*). Lack of such transparency in the design, development, and analysis of these models not only reduces the trust in their timely messages, but also limits the reproducibility of the models, impeding other scientists from verifying the findings and improving a model’s performance via further explorations and innovation. Many modelers have already shared the details of their models openly, yet the overall status of transparency of COVID-19 models remains unknown.

To systematically evaluate the transparency of COVID-19 models, we reviewed a sample of models that have earned global attention and been widely used to inform public health policies. We first collected COVID-19 models that included a methods write-up from CDC’s compilation (*3*), then collected the most-cited COVID-19 models in Google Scholar; this resulted in 29 models for evaluation. Due to the urgency of the pandemic, preprints and project websites made available in advance of publication and have had an essential role during the crisis (*4*), and therefore, we included models from these sources (n=12) in addition to peer-reviewed publications (n=17).

We assessed these sample models against 27 binary criteria to evaluate the transparency of their reports. Adopted from several transparency checklists (*5–7*) and tailored to evaluate models, two main themes guide the transparency assessment criteria: 1) specificity of model items, including but not limited to discussion of model mechanisms, assumptions, parametrization, formulation, codes, and sensitivity analysis; and 2) general research items, such as disclosure of research limitations, funding, and potential conflict of interest. Two trained researchers reviewed the full text and appendix of each modeling report and a third reviewer helped to discuss the discrepancies.

The results of our evaluation are reported in Figure 1. On average, the transparency criteria are satisfied by 75% of the sample models. While eight of the criteria are satisfied by 90% of the models, most of the criteria are satisfied by a much smaller percent of models. For instance, 21% of the models do not report the sources of their longitudinal data, 24% do not report their equations, 31% do not report their estimated parameters, 48% do not share their longitudinal data, and 52% do not report their code. Among the sample articles, only four articles satisfied 90% of our transparency checklist items.

**Fig. 1.**
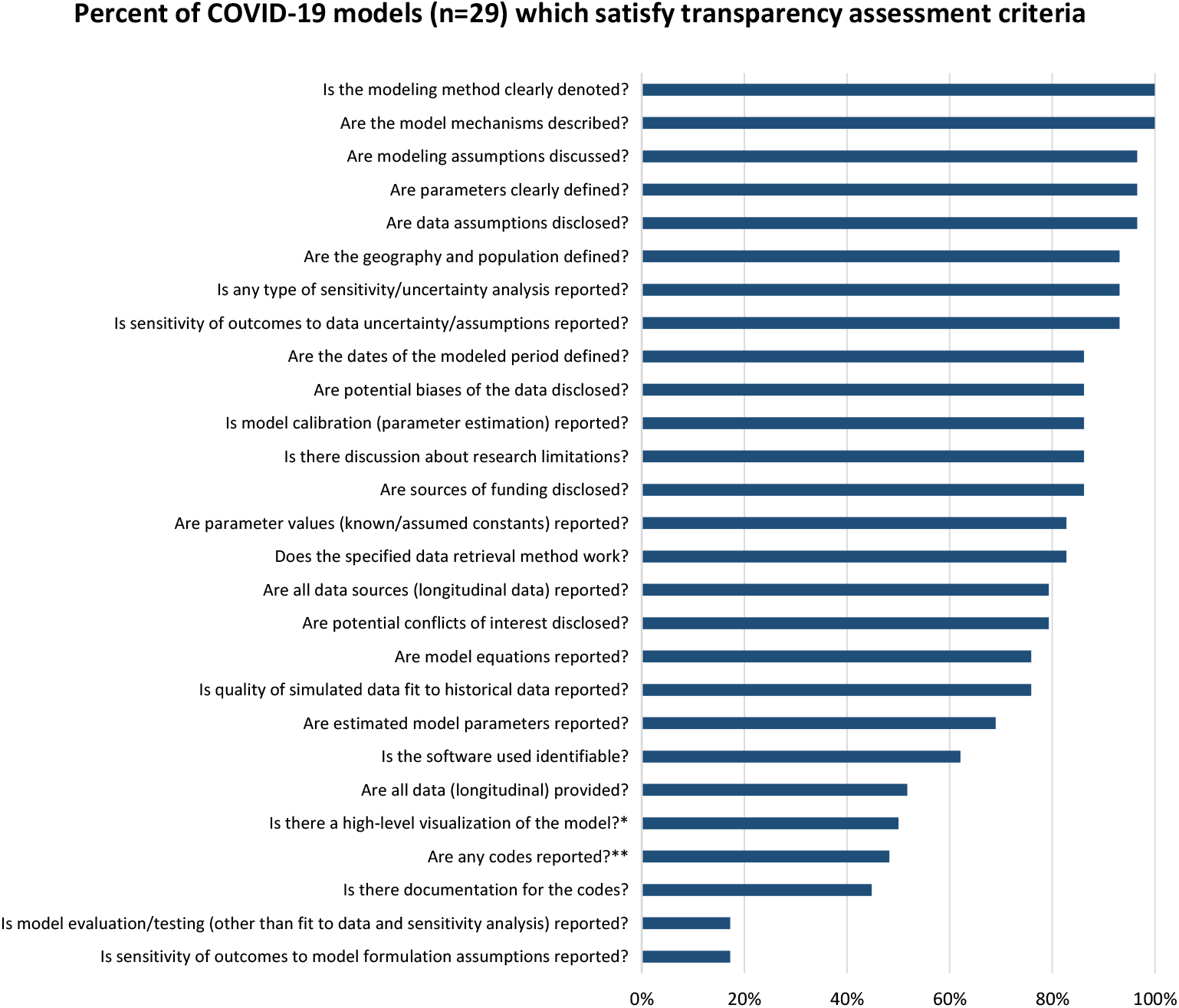
Percent of COVID-19 models (n=29) which satisfy transparency assessment criteria *Five regression models are exempt from this criterion, as there is no visualization that communicates model structure. **codes used for a generalized model are not sufficient; we were able to successfully retrieve the codes of each model that provided a retrieval method.

Evaluations like this one demonstrate that a model which is not fully transparent can still posit analytical insights and propose policy. Rather than presenting recommendations at face value, it is imperative for modelers to make sure that their claims are able to be independently verifiable. The scientific and modeling communities can and must hold themselves accountable to make transparency the norm, not the exception, or else risk losing the faith of policymakers and the public.

Such consequences were observed when IHME released their model in late March with highly criticized projections; they presented confidence intervals that converged in the future and seemingly low projections of case numbers and deaths during the first wave, among other things, and the scientific community was further frustrated by the lack of codes and other model details. This model was often cited by government agencies, including the White House (*8*), and its negative reception was difficult to overcome, even after their release of ostensibly more realistic projections. Another high-profile preprint model, published March 16, 2020 by Imperial College London, experienced similar scrutiny after predicting 510,000 COVID-19 deaths in a no-intervention scenario, prompting researchers to attempt replication. Unfortunately, the research team withheld their code for nearly six weeks following publication. Meanwhile, the United Kingdom instituted the recommended stringent stay-at-home measures. When the code was released in late April, several bugs and assumptions were unearthed and was not until June that the replication attempts were successful (*9*).

A crucial element of transparency is achieved by providing codes. One concern for researchers is that their code is disorganized and cannot be evaluated or run by laypeople hoping to replicate their efforts. However, this is a misconception; even messy code can provide a framework for an accurate replication and generate useful dialogue, as seen on platforms like GitHub. Still, well-documented code is preferable. Of the 48% of articles which reported their codes, all but one provided helpful detailed documentation either directly in the file or in a supplement. We encourage COVID-19 modelers who hope to impact perceptions and policy to release their codes in a timely manner for public evaluation.

Many journals ask for transparency statements and encourage scientists to report the details in supplemental documents. Research shows the data sharing policies of journals has increased the frequency and quality of data sharing altogether (*10*). While journals need to further enhance their publication policies and increase their transparency requirements, journals’ options are limited, and they cannot control the full transparency of publications. During a crisis such as COVID-19, preprints provide speedy information delivery before peer review, therefore, journal requirements and policies make minimal impact. Models which were still preprints or project websites satisfied an average of 70% of the transparency criteria, as compared to peer-reviewed articles’ 79%. The responsibility of transparency remains largely on the shoulders of modelers, even though the peer-review process can help address these omissions. It is imperative that modelers take it upon themselves to follow open research practices, adopt documentation and reporting guidelines, and share full details of their models.

Reporting a fully documented and transparent model can be difficult, but this effort has both tangible and intangible benefits for the modelers. With the urgency of a global pandemic, modelers might justify putting transparency second to the speed of reporting, however, poor transparency of models that directly impact public health policies and therefore human lives can have major harmful consequences. Hence, *all* models must be fully transparent for both scientific and ethical purposes.

## Data Availability

All data used in the assessment are reported in the supplementary document.

## Data and materials availability

All analysis details are available in the supplementary materials.

**Table S1:**
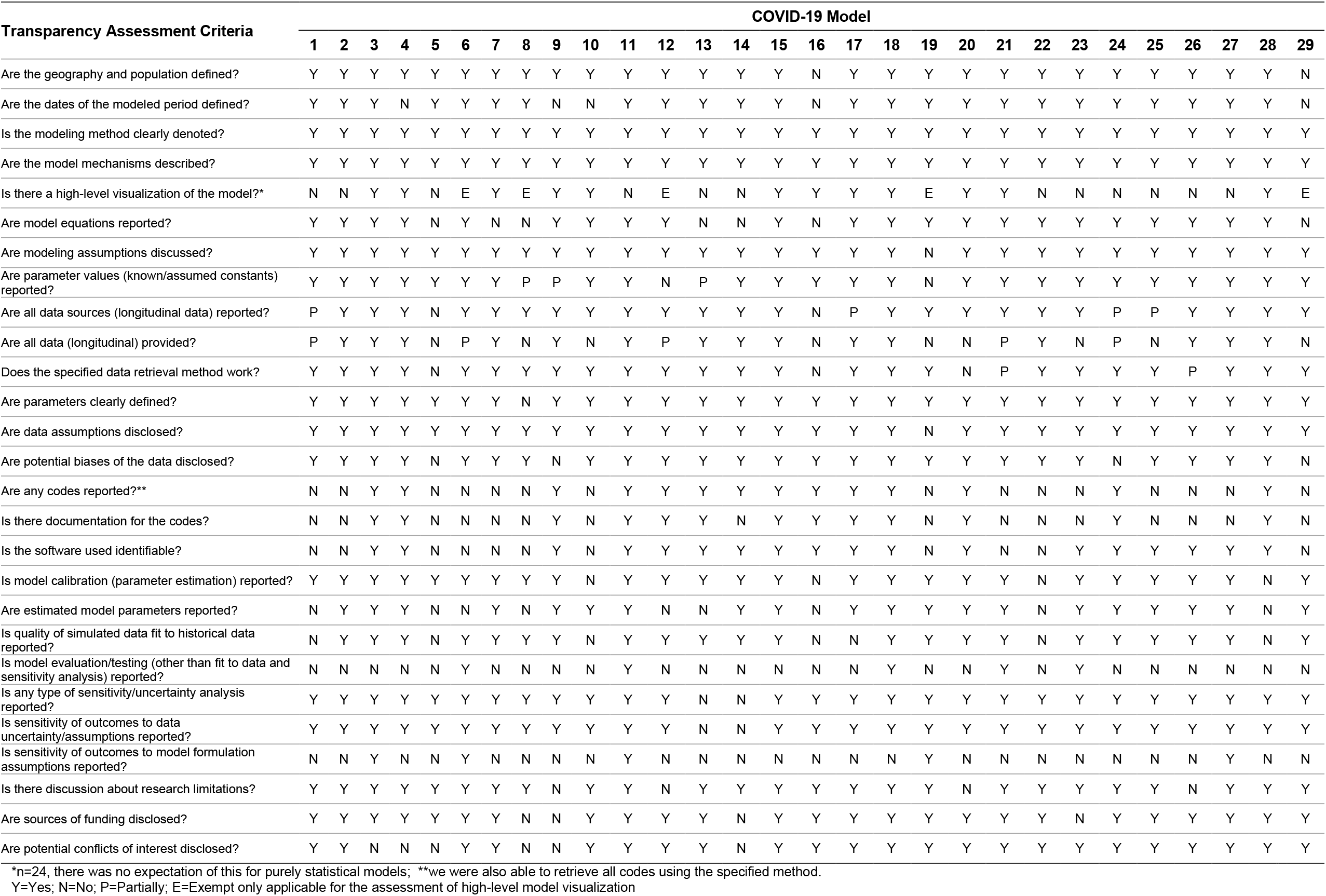
Transparency Assessment of 29 COVID-19 models

